# The impact of outdoor walking interventions on frailty among older adults with mobility limitations: findings from the Getting Older Adults Outdoors (GO-OUT) study

**DOI:** 10.1101/2025.04.19.25326113

**Authors:** Tai-Te Su, Ruth Barclay, Rahim Moineddin, Nancy M. Salbach

**Affiliations:** Department of Physical Therapy, University of Toronto, Toronto, Ontario, Canada; Department of Physical Therapy, University of Manitoba, Winnipeg, Manitoba, Canada; Department of Family and Community Medicine, University of Toronto, Toronto, Ontario, Canada; KITE Research Institute, Toronto Rehabilitation Institute - University Health Network, Toronto, Ontario, Canada

## Abstract

**Objective:** Diverse strategies are needed to reduce frailty. This study evaluated the effects of two behavioural interventions targeting outdoor walking on reducing the level of frailty among community-dwelling older adults with mobility limitations.

**Methods:** Data from two participant cohorts of the Getting Older Adults Outdoors (GO-OUT) study were analyzed. After baseline evaluations, 190 participants were invited to a one-day educational workshop and were then randomized to either a 10-week supervised outdoor walk group (n=98) or a 10-week telephone weekly reminders group (n=92). We assessed frailty using Fried’s frailty index at 0, 3, and 5.5 months. Mixed-effects linear and ordinal regression models were used to evaluate change in frailty score and phenotype over time after accounting for age, sex, study site, participation on own or with a partner, and cohort.

**Results:** At baseline, participant mean age was 74.5 ± 7.1 years; 73% were female, 7% were frail, and 59% were pre-frail. Total frailty scores decreased, on average, by 0.13 points (*b* = – 0.13, 95% CI: –0.26 to –0.01; *p*=.036) across all participants from 0 to 3 months (immediately post-intervention). Participants were 55% less likely to progress to more severe frailty phenotypes at 3 months compared to baseline (OR=0.45; 95% CI: 0.25 to 0.81; *p*=.008). No significant between-group differences or long-term effects were observed.

**Conclusions:** A short-term reduction in frailty was observed in older adults with mobility limitations following participation in behavioural interventions aimed at improving outdoor walking; neither intervention was superior. Supervised outdoor walk group and telephone weekly reminder interventions to increase outdoor walking may have the potential to mitigate frailty in older adults with mobility limitations.

## Introduction

Frailty is a clinical syndrome characterized by age-related decline in physiological reserve and function, reflecting a state of vulnerability to stressors and poor health outcomes [1–4]. Frailty is commonly seen in older adults [5]. In Canada, for instance, the prevalence of frailty is 5.3% among adults aged 18-34 years and increases to 7.8% among older adults aged 65 years and older [6]. While various approaches have been developed to assess frailty [2,3,7], the Fried’s frailty index is one of the most frequently adopted metrics in both research and clinical settings [2,8,9]. According to Fried’s model, frailty is operationally defined as physical phenotypes consisting of five indicators: unintentional weight loss, exhaustion, low physical activity, muscle weakness, and slow walking speed [1]. Individuals are classified as non-frail (0 indicators present), pre-frail (1 or 2 indicators present), or frail (3 or more indicators present). Importantly, frailty has been linked to adverse health outcomes such as falls, institutionalization, hospitalization, and premature mortality [10–12]. Research has also shown, however, that frailty is dynamic and reversable in nature, even in later life, highlighting opportunities for intervention efforts to delay or attenuate frailty among older adults [13–15].

Exercise plays a pivotal role in preventing and managing frailty. A growing body of evidence demonstrates that interventions involving aerobic, resistance, balance, and flexibility training can improve physical performance and mitigate frailty among community-dwelling older adults and those residing in long-term care facilities [16–19]. Notably, walking and jogging were rated as the most popular forms of exercise in a nationally representative sample of older adults in the United States [20]. Walking, especially in an outdoor environment, has shown particular promise in addressing frailty. For example, outdoor Nordic walking has been found to be more effective than general exercise in improving lower extremity strength, reducing weakness, and alleviating depression in frail older adults aged 70 years and older [21]. Although the overall benefits of exercise interventions are well-documented, prior research reveals that these programs are often insufficient to stimulate or sustain increases in physical activity both during and after the intervention period [22,23]. Given that physical inactivity is a known risk factor for frailty, it is essential to develop broader intervention strategies that promote long-term behaviour change. These strategies could focus on promoting healthy lifestyles and habits, reducing sedentary behaviour, and supporting continued physical activity in everyday life [24,25].

The Getting Older Adults Outdoor (GO-OUT) study [26] offers an exceptional opportunity to examine the effects of two behavioural interventions on improving frailty outcomes. Conducted across four large cities in Canada, the GO-OUT study compared a 1-day educational workshop followed by a 10-week, park-based, supervised, outdoor walk group (OWG) program, to the same workshop and a 10-week weekly reminders (WR) program, in promoting outdoor walking activity among older adults with mobility limitations [27]. The GO-OUT interventions were designed *a priori* to improve outdoor walking [26], a key indicator of physical function and a central element in frailty assessment. Both GO-OUT interventions have the potential to enhance older adults’ physical health and may inform the development of frailty mitigation and prevention strategies. The aim of this study was to evaluate whether a 10-week OWG program or a 10-week WR program could reduce the level of frailty in community-dwelling older adults with mobility limitations. We hypothesized that each intervention would be beneficial, with participants in the OWG program more likely to show a greater reduction in frailty indicators and a lower risk of progressing to worse frailty phenotypes compared to their baseline measures and participants in the WR program.

## Methods

### Study design and procedures

The GO-OUT study was a four-site, two-parallel-group randomized trial conducted in Edmonton, Winnipeg, Toronto, and Montreal, Canada. The GO-OUT study was registered on ClinicalTrials.gov (registration number: NCT03292510). Between February 20, 2018 and May 15, 2019, two cohorts of older adults who met the eligibility criteria detailed below were enrolled and invited to participate in a 1-day educational workshop. Immediately following the workshop, the research personnel stratified the190 participants by study site and type (enrolled as an individual or with a partner) and randomized them into either a 10-week outdoor walk group (OWG; n=98) or a 10-week weekly reminders group (WR; n=92). The random allocation sequence was computer-generated by a biostatistician external to the research team and implemented in REDCap [28]. The study protocol received approval from the research ethics broads at each study site [26]. The current study was conducted with approval from the University of Toronto’s research ethics board (Protocol #35251).

### Participants

To be eligible for the GO-OUT study, participants needed to be aged 65 years or older, reporting difficulty walking outdoors, and living independently in the community. They were required to have the ability to walk at least one block (∼50 meters) independently, with or without a walking aid, express a willingness to sign a liability waiver or obtain physician clearance for exercise, exhibit mental competency by scoring at least 18 out of 22 on the Mini-Mental State Examination (telephone version) [29], be available to participate in the workshop and at least five weeks of the OWG program, and be able to speak and understand English. Individuals were excluded if they self-reported engaging in physical activity for 150 minutes or more per week, were currently receiving rehabilitation to improve walking, or were classified as at a high risk for falls [30]. We gave the option to participate with a partner. All participants signed a written informed consent form prior to baseline evaluation.

### Data collection

Trained evaluators who were masked to the intervention assignment conducted evaluations at baseline (0 months), 3 months, 5.5 months, and 12 months. Frailty was assessed using the Cardiovascular Health Study Frailty Index [1]. We determined the presence of five frailty indicators: 1) weight loss; 2) exhaustion; 3) low physical activity; 4) weakness; and 5) slowness. *Weight loss* was measured using the question “In the last year, have you lost more than or equal to 10 lbs or 4.5 kg unintentionally (i.e. not due to dieting or exercise)?” This indicator was present if the participant answered yes. *Exhaustion* was measured using two statements from the Center for Epidemiological Studies–Depression (CES-D) scale [31], including (a) “I felt that everything I did was an effort” and (b) “I could not get going”. Participants were asked to rate how often they felt this way over the past week for each statement. The exhaustion criterion was met if participants reported feeling this way a moderate amount of the time (3–4 days) or most of the time for either statement. *Low physical activity* was assessed using the short version of the Minnesota Leisure Time Activity questionnaire [32]. Participants reported the amount of time spent on the following 15 activities during the past two weeks: walking for exercise, moderately strenuous chores, mowing the lawn, raking the lawn, gardening, hiking, jogging, biking, exercise cycle, dancing, aerobics/aerobic dance, bowling, golf, calisthenics, and swimming. Participants could specify up to two additional activities. We calculated total energy expenditure in kilocalories (kcals) per week using standardized activity codes and algorithms [33]. Low physical activity was defined as total energy expenditure below the gender-specific threshold [1]. *Weakness* was measured using grip strength and was considered present if participants’ average grip strength across three trials was lower than the cut-off values stratified by gender and body mass index (BMI) quartiles [1]. *Slowness* was originally determined by the time taken to walk 15 feet. Since the GO-OUT study used the 10-meter walk test [26], we converted the units to ensure consistency in the calculations. The slowness indicator was deemed present if participant’s walk time exceeded the cut-off values stratified by gender and height [1]. Presence of an indicator was assigned a score of 1 point. Scores were summed and could range from 0 (no indicators present) to 5 (5 indicators present). In addition, based on Fried’s criteria, participants were classified into one of three frailty phenotypes: *non-frail* (0 indicators), *pre-frail* (1–2 indicators), and *frail* (3–5 indicators). Due to the COVID-19 pandemic, we were unable to administer performance-based measures, such as grip strength and walking speed, to measure frailty among participants in Cohort 2 at the 12-month evaluation. Consequently, frailty data collected at baseline, 3 months, and 5.5 months were analyzed in this study. S1 Table presents the percentage of participants with missing frailty score by intervention group and time point.

Information on age (years), sex (female vs. male), body mass index (BMI), educational attainment, number of comorbidities, and physical capacity (balance, walking endurance, walking speed, etc.) was collected at baseline to describe participants’ characteristics.

### Educational workshop

All participants were scheduled for a 1-day, interactive educational workshop before randomization. The workshop aimed to enhance knowledge, self-efficacy, and skills to engage in outdoor walking. Participants circulated in small groups through eight stations, including 1) Canadian physical activity guidelines for older adults; 2) setting SMART (specific, measurable, achievable, realistic, and timely) goals; 3) pedometer use; 4) Nordic pole walking; 5) footwear, footcare, and proper walking patterns; 6) fall prevention; 7) monitoring exercise intensity and safety; and 8) balance exercises and postural awareness. Each station was facilitated by a student in a health-related program or a health professional with experience working with older adults and individuals living with chronic conditions. All participants received a pedometer and a workbook with content and tools reviewed during the workshop.

### Outdoor walk group program

The OWG program was developed based on a conceptual framework of community mobility [34]. The OWG program aimed to improve older adults’ competency in multiple dimensions of community mobility such as walking distance, walking speed, postural transitions, attentional demands, and ambient conditions [34]. The OWG consisted of two 1-hour walking sessions each week for a duration of 10 weeks, resulting in a maximum of 20 sessions. Each session was group-based, held in large neighborhood parks, and structured to include a 10-minute warm-up, a continuous distance walk, task-oriented practice of an outdoor walking skill, a second continuous distance walk, and a 10-minute cool-down. The program was implemented during summer months from June to August, with progressively increasing walking distances and different walking activities each week. Sessions were led by a health professional (e.g., physiotherapist or kinesiologist) who had completed a 90-minute training session and were authorized to modify the program’s difficulty level to ensure optimal challenge for participants. Group leaders received assistance from OWG assistants to maintain a facilitator-to-participant ratio of 1:3, with a maximum of three facilitators and nine participants per group.

### Weekly reminders

The WR intervention was implemented to facilitate outdoor walking and ensure that participants in each group received attention throughout the intervention period. Following the educational workshop, participants assigned to the WR group received a phone call from study coordinators every week for a duration of 10 weeks. Each reminder followed a structured script designed to encourage participants to review and discuss the content delivered during the workshop, including getting physically active, setting SMART goals, home balance exercises, and prevention of falls. In situations where participants could not be reached by phone, scripted email reminders were sent as an alternative.

### Statistical analysis

Differences in baseline characteristics between the OWG and WR group were assessed using two-sample *t* test and chi-squared test. To characterize participants’ frailty status, we reported frailty sum scores with means and standard deviations (SDs) and frailty phenotypes with counts and percentages. We calculated the numbers and rates of transitions between three frailty phenotypes across each evaluation time point. In situations where participants had missing data for one to four of the five frailty indicators (n=19 at baseline), we followed established recommendations and allowed only one missing indicator when a person had a frailty sum score between 0 and 2. For those with a frailty sum score of 3 or higher, up to two missing indicators were allowed to reduce misclassification of the frailty phenotype [35]. This approach applies stricter criteria to account for missing data compared to earlier methods, where participants with two or fewer missing indicators were considered evaluable for frailty [1].

Mixed-effects regression models, which account for the repeated observations within individuals, were used to estimate change in frailty status from baseline to 3 months and baseline to 5.5 months. We adopted a linear mixed-effects model for frailty sum scores and a mixed-effects ordinal regression model for frailty phenotypes. For both frailty outcomes, we first tested the fixed effects of intervention group, time, and their interaction. As the interaction term was not statistically significant, it was removed from subsequent models. Visual inspection of frailty trajectories confirmed that changes were comparable between the OWG and WR group over time (please see S1 Fig). Next, we adjusted for potential confounders, including participant age and sex, and accounted for study site, participant type, and cohort reflecting the complex design of the GO-OUT study. Random effects for time of assessment were tested but did not improve model fit and were excluded. We assessed model assumptions prior to interpretation of results. Recognizing the small proportion of frail participants (<10%) in the present study, we conducted sensitivity analysis using mixed-effects logistic regression models with a broader definition of frailty (i.e., combining pre-frail and frail participants versus non-frail participants) to assess the robustness of our findings. A two-tailed *p*-value <.05 was deemed statistically significant. We performed the analyses in R (version 4.1.0) using the lmer and ordinal packages [36,37].

## Results

Table 1 summarizes baseline characteristics of all GO-OUT participants and by intervention group. The mean age was 74.5 years (SD ±7.1) among the pooled sample, and the majority of participants were female (73%). Forty-four percent of the participants had obtained a bachelor’s degree or higher. Individual characteristics were similar between the two intervention groups at baseline (all *p*-values>.05). Regarding intervention adherence, participants in the OWG attended a median of 13 sessions (interquartile range [IQR]: 7–17) out of 20. Participants in the WR group received a median of 9 reminders (IQR: 9–10) out of 10 over the 10-week intervention period.

**Table 1.** Participant characteristics at baseline by GO-OUT intervention group.

| Characteristic | Pooled<br>(n = 190) | Outdoor walk group<br>(n = 98) | Weekly reminders<br>(n = 92) | p-value |
| --- | --- | --- | --- | --- |
| | Mean ± SD / Number (%) | | | |
| Study site |  |  |  | .99 |
| Site 1 | 51 (27) | 26 (27) | 25 (27) |  |
| Site 2 | 53 (28) | 28 (29) | 25 (27) |  |
| Site 3 | 50 (26) | 26 (27) | 24 (26) |  |
| Site 4 | 36 (19) | 18 (18) | 18 (20) |  |
| Participant type |  |  |  | .73 |
| Individual | 154 (81) | 78 (80) | 76 (83) |  |
| Dyad | 36 (19) | 20 (20) | 16 (17) |  |
| Cohort |  |  |  | .99 |
| 2018–19 | 65 (34) | 33 (34) | 32 (35) |  |
| 2019–20 | 125 (66) | 65 (66) | 60 (65) |  |
| Age (years) | 74.5 ± 7.1 | 75.3 ± 6.9 | 74.2 ± 7.4 | .58 |
| 65–74 years | 106 (56) | 54 (55) | 52 (57) | .86 |
| 75–84 years | 59 (31) | 32 (33) | 27 (29) |  |
| 85+ years | 25 (13) | 12 (12) | 13 (14) |  |
| Sex |  |  |  | .55 |
| Female | 139 (73) | 74 (76) | 65 (71) |  |
| Male | 51 (27) | 24 (24) | 27 (29) |  |
| Body mass index (kg/m<sup>2</sup>) | 29.5 ± 6.3 | 29.6 ± 6.4 | 29.3 ± 6.2 | .77 |
| Educational attainment |  |  |  | .93 |
| Secondary or lower | 39 (21) | 21 (21) | 18 (20) |  |
| Some or completed college | 68 (36) | 34 (35) | 34 (37) |  |
| Bachelor's degree or higher | 83 (44) | 43 (44) | 40 (43) |  |
| Uses a walking aid | 48 (25) | 28 (29) | 20 (22) | .36 |
| Charlson comorbidity index (0–39) | 1.99 ± 1.91 | 2.05 ± 2.01 | 1.93 ± 1.80 | .68 |
| 6-minute walk test (m) | 358.5 ± 91.2 | 359.0 ± 88.5 | 357.9 ± 94.3 | .94 |
| 10-meter walk test comfortable (m/s) | 1.07 ± 0.23 | 1.08 ± 0.24 | 1.06 ± 0.22 | .69 |
| 10-meter walk test fast (m/s) | 1.41 ± 0.32 | 1.41 ± 0.31 | 1.41 ± 0.32 | .91 |
| Mini-BESTest (0–28) | 20.4 ± 5.2 | 20.2 ± 5.0 | 20.6 ± 5.5 | .60 |
| 30-second sit-to-stand (count) | 8.3 ± 4.1 | 8.1 ± 3.7 | 8.6 ± 4.4 | .32 |
| ASCQ (0–10)† | 7.9 ± 1.6 | 7.8 ± 1.7 | 8.0 ± 1.5 | .28 |
| RAND-36 well-being (0–100) | 75.9 ± 15.5 | 75.4 ± 16.9 | 76.3 ± 14.0 | .70 |

Note: † ASCQ = ambulatory self-confidence questionnaire. SD = Standard deviation. p-values indicate the statistical significance of differences in baseline participant characteristics between the outdoor walk group (OWG) and the weekly reminders (WR) group.

Fig 1 presents the number of participants at each time point. Approximately 80% of participants remained in the study at 3 months, and 70% remained at 5.5 months. Retention rates were similar across intervention groups. Baseline characteristics of study completers and withdrawals are provided in S2 Table. Multivariable logistic regression analyses showed that withdrawal was more prominent among participants from site 1, those enrolled in the 2019-20 cohort, and those with a higher BMI. Intervention group and frailty status were not significant predictors of withdrawal.

**Fig 1.**
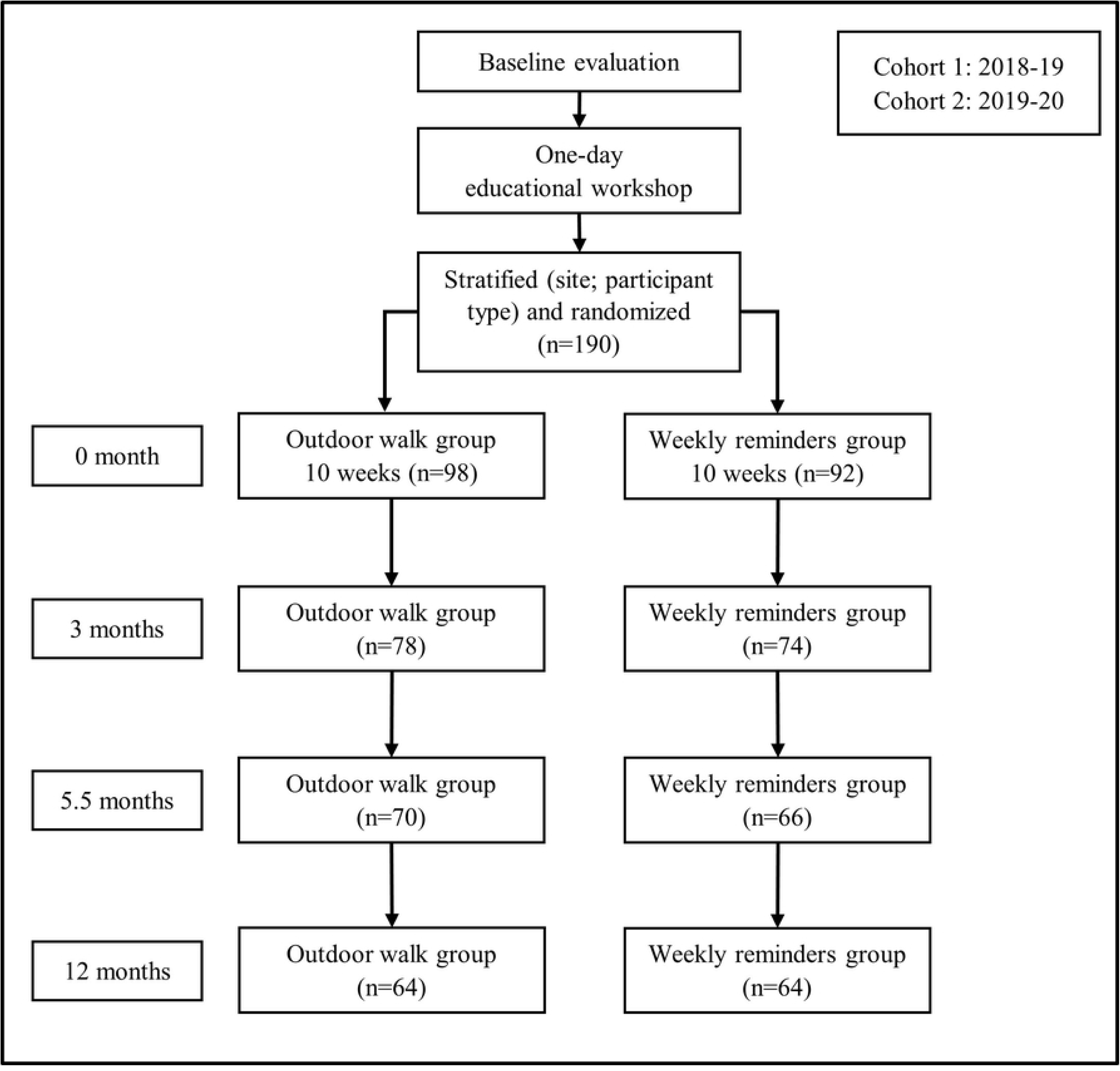
Illustration of the GO-OUT study design and number of participants at each evaluation time point *Note*: Due to COVID-19 precautions, we were unable to collect performance-based outcome measures for frailty assessment, including grip strength and walking speed, from participants in Cohort 2 at the 12-month time point. Hence, we focused on data collected at baseline, 3 months, and 5.5 months for the present study. Detailed information on missing frailty scores at each time point is provided in S1 Table.

Table 2 summarizes frailty status at each time point. The mean frailty sum scores (out of 5) were 0.98±0.91 at baseline, 0.82±0.91 at 3 months, and 0.95±0.99 at 5.5 months. At baseline, 34% of participants were classified as non-frail, 59% as pre-frail, and 7% as frail. By 3 months, the percentage of non-frail participants increased to 45%, and the percentages of pre-frail and frail participants decreased to 49% and 5%, respectively. However, by 5.5 months, the percentage of non-frail participants declined to 40%, with pre-frail and frail participants rising to 51% and 9%. No significant differences in frailty sum scores or phenotypes were found between the OWG and WR group at each time point.

**Table 2.** Comparison of frailty status between intervention groups at each evaluation time point.

| Frailty measure | Pooled | Outdoor walk group | Weekly reminders |  |
| --- | --- | --- | --- | --- |
| | Mean ± SD / Number (%) | | | *p*-value |
| Frailty score (0–5) |  |  |  |  |
| Baseline | 0.98 ± 0.91 | 1.02 ± 0.94 | 0.93 ± 0.87 | .51 |
| 3 months | 0.82 ± 0.91 | 0.88 ± 0.96 | 0.75 ± 0.86 | .38 |
| 5.5 months | 0.95 ± 0.99 | 1.02 ± 0.99 | 0.89 ± 0.99 | .47 |
| Frailty phenotype |  |  |  |  |
| Baseline |  |  |  | .36 |
| Non-frail (0) | 63 (34) | 30 (31) | 33 (37) |  |
| Pre-frail (1–2) | 110 (59) | 57 (59) | 53 (59) |  |
| Frail ($\geq 3$) | 13 (7) | 9 (9) | 4 (4) | |
| 3 months |  |  |  | .77 |
| Non-frail (0) | 66 (45) | 32 (43) | 34 (48) |  |
| Pre-frail (1–2) | 72 (49) | 38 (51) | 34 (48) |  |
| Frail ($\geq 3$) | 8 (5) | 5 (7) | 3 (4) | |
| 5.5 months |  |  |  | .38 |
| Non-frail (0) | 51 (40) | 23 (35) | 28 (44) |  |
| Pre-frail (1–2) | 65 (51) | 34 (52) | 31 (49) |  |
| Frail ($\geq 3$) | 12 (9) | 8 (12) | 4 (6) | |

Note: *p*-values indicate the statistical significance of differences in frailty status between the outdoor walk group (OWG) and the weekly reminders (WR) group at each evaluation time point.

Table 3 presents the number and rate of transitions between the three frailty phenotypes at each time point. From baseline to 3 months, 52.7% of participants maintained their frailty status, 16.1% improved, and 8.1% transitioned to a more severe state. Between 3 and 5.5 months, 53.4% of participants remained stable, but improvements dropped to 8.9%, while worsening transitions increased to 20.5%. The transition patterns from baseline to 3 months were similar in the OWG and WR group, where around 70% participants remained stable or showed improvement. Yet, from 3 to 5.5 months, 54.7% of participants in the OWG remained stable or improved, compared to 70.5% in the WR group. Despite observing various transitions, no participants in either group transitioned from frail to non-frail or vice versa during the follow-up period. Details on the presence of the five frailty indicators are presented in S3 Table. Notably, the proportion of OWG participants with the weakness indicator decreased from 47% at baseline to 42% at 3 months, and the slowness indicator decreased from 5% to 0%. In the WR group, the proportion with the weakness indicator decreased from 36% at baseline to 28% at 3 months, while the low activity indicator dropped from 15% to 10% over this time period.

**Table 3.**
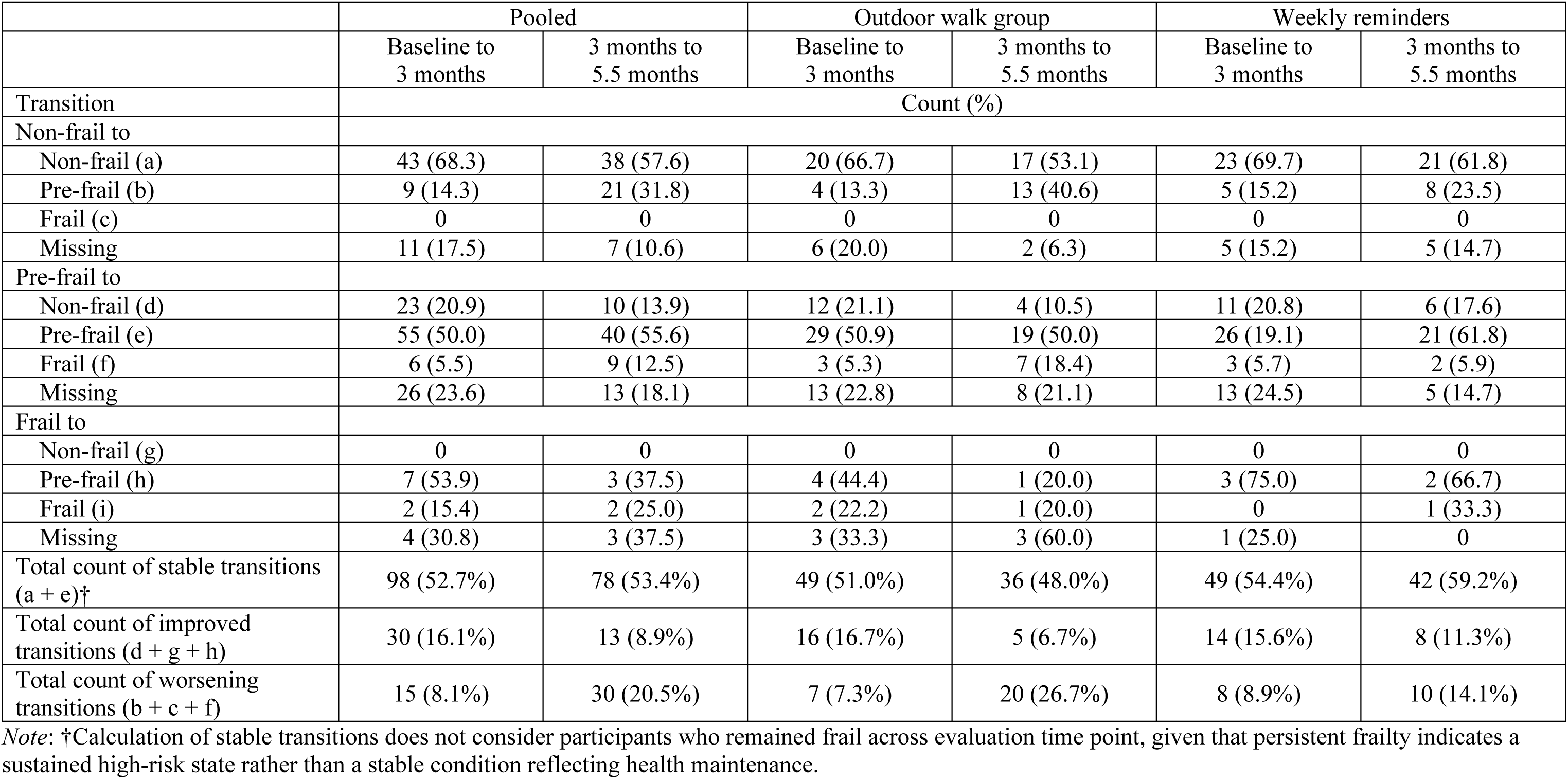
Numbers and rates of transitions between frailty phenotypes across each evaluation time point.

Table 4 presents results from the mixed-effects models examining changes in frailty status over time. In the unadjusted model, Fried’s frailty sum scores decreased by an average of 0.13 points (*b* = –0.13, 95% CI: –0.26 to –0.00; *p* = .043) across participants in both OWG and WR group from baseline to 3 months. Similarly, the odds of experiencing greater frailty (i.e., being pre-frail or frail versus non-frail) were 55% lower (OR=0.45; 95% CI: 0.25, 0.82; *p*=.009) among all participants at 3 months compared to baseline. However, there were no significant changes in frailty status from baseline to 5.5 months. Additionally, differences in either frailty sum scores or phenotypes between the OWG and WR group were not statistically significant. These results remained consistent after accounting for participants age, sex, and the nested study design features, and were robust in the sensitivity analysis, which grouped pre-frail and frail participants together versus non-frail participants (S4 Table). S1 Fig 1 and S2 Fig visually illustrate changes in frailty scores and phenotypes across each time point.

**Table 4.** Results of mixed-effects models of change in frailty over time in the pooled sample of GO-OUT participants.

|  | Outcome 1<br>Frailty sum scores (0–5) |  |  |  | Outcome 2<br>Frailty phenotypes (non-frail; pre-frail; frail) |  |  |  |
| --- | --- | --- | --- | --- | --- | --- | --- | --- |
|  | Model 1 |  | Model 2 |  | Model 1 |  | Model 2 |  |
| Variables | *Beta* [95% CI] | *p* | *Beta* [95% CI] | *p* | OR [95% CI] | *p* | OR [95% CI] | *p* |
| Time |  |  |  |  |  |  |  |  |
| Baseline | *Reference* |  | *Reference* |  | *Reference* |  | *Reference* |  |
| 3 months | −0.13 [−0.26, −0.00] | .043 | −0.13 [−0.26, −0.01] | .036 | 0.45 [0.25, 0.82] | .009 | 0.45 [0.25, 0.81] | .008 |
| 5.5 months | 0.04 [−0.09, 0.18] | .521 | 0.03 [−0.10, 0.16] | .628 | 1.02 [0.55, 1.88] | .950 | 0.97 [0.54, 1.77] | .933 |
| Intervention |  |  |  |  |  |  |  |  |
| WR | *Reference* |  | *Reference* |  | *Reference* |  | *Reference* |  |
| OWG | 0.14 [−0.10, 0.38] | .266 | 0.14 [−0.08, 0.36] | .212 | 2.26 [0.82, 6.22] | .116 | 2.26 [0.93, 5.46] | .071 |
| Age† | — |  | 0.02 [0.00, 0.04] | .027 | — |  | 1.07 [1.00, 1.15] | .038 |
| Sex | — |  |  |  | — |  |  |  |
| Male |  |  | *Reference* |  |  |  | *Reference* |  |
| Female |  |  | −0.39 [−0.64, −0.14] | .002 |  |  | 0.16 [0.06, 0.45] | <.001 |
| Type | — |  |  |  | — |  |  |  |
| Individual |  |  | *Reference* |  |  |  | *Reference* |  |
| Dyad |  |  | −0.08 [−0.37, 0.21] | .587 |  |  | 0.66 [0.21, 2.08] | .479 |
| Cohort | — |  |  |  | — |  |  |  |
| 2018-19 |  |  | *Reference* |  |  |  | *Reference* |  |
| 2019-20 |  |  | −0.11 [−0.34, 0.13] | .369 |  |  | 0.67 [0.26, 1.69] | .392 |
| Study site | — |  |  |  | — |  |  |  |
| Site 1 |  |  | *Reference* |  |  |  | *Reference* |  |
| Site 2 |  |  | 0.27 [−0.07, 0.61] | .119 |  |  | 5.24 [1.31, 20.97] | .019 |
| Site 3 |  |  | 0.62 [0.30, 0.94] | <.001 |  |  | 12.18 [3.15, 47.01] | <.001 |
| Site 4 |  |  | 0.19 [−0.10, 0.49] | .201 |  |  | 2.26 [0.67, 7.59] | .187 |
| Variance ($\sigma^2$) | | | | | | | | |
| Intercept ($U_{0i}$) | 0.56 | | 0.43 | | 8.94 | | 5.90 | |
| Residuals ($e$) | 0.31 | | 0.31 | | — | | — | |

Note: WR = Weekly reminders. OWG = Outdoor walk group. OR = Odds ratio. CIs = Confidence intervals. Linear mixed-effects models and mixed-effects ordinal regression were used to estimate changes in frailty sum scores (outcome 1) and frailty phenotypes (outcome 2), respectively. Model 2 controlled for participant age, sex, and study clustering variables including study site, participant type, and cohort. †Age was centered at 63 years, representing the minimum age among the study participants.

## Discussion

To our knowledge, this study is among the first to evaluate the effects of two distinct interventions on frailty, with one involving outdoor walking and the other using telephone reminders, among community-dwelling older adults with mobility limitations. Results indicate that participants in both the supervised, group-based OWG and the telephone-based WR group experienced similar reductions in frailty scores from baseline to 3 months. In addition, although no between-group differences were found, participants in both programs were less likely to progress toward more severe frailty phenotypes immediately post-intervention.

Overall, our findings show that there was a short-term improvement in frailty among the GO-OUT study participants as a whole. The absence of significant group effects suggests that no single intervention is superior to the other, and that both the OWG and WR programs may have contributed to the observed improvement. To start with, the use of physical exercise as a therapy for frailty is well-established, with growing evidence supporting its effectiveness [18,38–41]. For instance, a meta-analysis of 1,068 older adults (ages 75.3 to 86.8) found that physical exercise significantly improved gait speed, balance, and performance in activities of daily living in frail older adults [42]. Among various forms of physical exercise, walking is considered the most feasible and accessible and is closely linked to life-space mobility, a key indicator of the area a person moves through and the development of clinical frailty [38,40,43,44]. In particular, a seminal theoretical framework derived from the Women’s Health and Aging Studies (WHAS) identifies constricted life-space as a marker of declining physiological reserve, leading to decreased physical activity, social isolation, and accelerated deconditioning, which in turn contribute to disability, frailty, and mortality in older adulthood [45]. The present study adds to the literature by showing that outdoor walking, which was practiced in the OWG and encouraged in the WR group, may help improve frailty among older adults with mobility limitations. As the OWG program was designed specifically to build individuals’ skills and self-efficacy to walk outdoors in groups, this approach holds promise for enhancing walking capacity, breaking the inactivity-frailty cycle, and fostering social engagement among older adults [41]. Importantly, our pilot study and quantitative process evaluation showed that OWG programs can be safely implemented in the community, with flexible adjustments made to accommodate participants’ varying abilities [46,47]. These findings suggest that a supervised, group-based outdoor walking program may be a promising option for improving frailty in later life.

Meanwhile, changing lifestyle habits represents an ongoing challenge for older adults who are frail or at risk of frailty. Despite the well-known benefits of physical exercise, prior research indicates that physical activity levels may not necessarily increase as a consequence of an exercise intervention [22]. In this regard, behavioural or reminder-based approaches may offer viable alternatives. In this study, participants in the WR group showed similar reductions in frailty scores as those in the OWG from baseline to 3 months. This result is unsurprising given the nature of the reminders offered, where participants received consistent prompting to walk outdoors, reviewed physical activity guidelines, and used self-monitoring technologies such as pedometers. Previous research has demonstrated that tailored advice, supplemented with written materials and information on local exercise facilities, can be as effective as supervised exercise programs in increasing physical activity [48]. Moreover, a randomized trial by Olaya-Contreras and colleagues found that tailored advice can promote active behaviours, as evidenced by increased daily step count, in patients with acute severe low back pain [49]. More broadly, reminder-based interventions have also been shown to improve medication adherence and behavioural/lifestyle modifications in people with various chronic conditions [25,50]. It is worth mentioning that the improvements seen in the WR group may reflect both the effect of the reminder itself and the potential impact of the Hawthorne effect, where participants alter their behaviour due to the awareness of being observed [51]. Nonetheless, our findings suggest that a reminder-based, minimally-supervised, and low-cost intervention may improve frailty outcomes and should be considered for individuals who prefer to exercise independently or when conventional exercise classes are not feasible due to resource constraints.

Contrary to our hypothesis, there were no significant differences in changes in frailty between the OWG and WR group from baseline to 3 months or baseline to 5.5 months. Our findings align with a recent systematic review and meta-analysis showing no significant advantage of physical activity interventions over usual care in improving frailty [52]. Several factors may shed light on this phenomenon. First and foremost, mean attendance rates differed markedly in the present study, with 63.1% for the OWG compared to 90.0% for the WR group. OWG participants reported barriers such as feeling unwell, transportation difficulties, employment, and other commitments as reasons for missing their outdoor walking sessions [27]. Lower attendance among the OWG participants directly influenced the amount of training they received and may thus explain the lack of differences in changes in frailty outcomes compared to the WR group. Second, we noted distinct patterns of improvements in the five frailty indicators between the two groups (S3 Table). In the OWG, a 5% reduction in the weakness and slowness indicators and a 2% reduction in the low activity indicator were observed from baseline to 3 months. Meanwhile, the WR group showed an 8% reduction in the weakness indicator, followed by a 5% reduction in the low activity indicator over the same period. These descriptive results suggest a divergence in the patterns of change and reflect the task-specificity of the two interventions: the OWG provided structured opportunities for older adults to practice outdoor walking tasks in large parks, while the WR encouraged participants to integrate physical activity into their daily routines. From a clinical perspective, each program addressed different aspects of frailty. Although the overall changes in the frailty score appear similar, the OWG and WR programs likely achieved these results through distinct mechanisms, highlighting the different pathways through which interventions can influence frailty outcomes. Notably, no significant changes in frailty were observed from baseline to 5.5 months in either the OWG or WR group.

We speculate that participants may have reduced their engagement in physical activity after the conclusion of the structured GO-OUT study. Additionally, the 5.5-month evaluation took place during Canada’s cold-weather months (October and November), and over half of the participants in both groups reported not walking outdoors during this period [27]. The seasonal effect may have limited the interventions’ ability to sustain improvements in frailty. These findings underscore both the potential and challenges of designing, implementing, and evaluating frailty interventions in real-world settings.

Our study offers two practical programs for consideration in frailty research and clinical practice. Each program targets different components of frailty and emphasizes strategies that are feasible and adaptable. Given the lack of a one-size-fits-all solution to frailty [53], our findings underscore the importance of providing diverse approaches to address frailty in aging populations. However, study limitations should be acknowledged. First, the COVID-19 pandemic impacted participant recruitment and disrupted frailty data collection, potentially limiting our ability to detect greater changes or evaluate long-term effects (12 months). Second, some participants in both groups reported receiving co-interventions (e.g., physical therapy or other exercise programs) during the study period, making it challenging to solely attribute changes to the interventions under investigation [27]. Third, while the Fried’s frailty index is a widely used measure, the absence of a minimal clinically important difference (MCID) value hindered interpretation of the observed improvements [54]. The mobility-focused nature of both the OWG and WR programs may have limited their ability to address other aspects of frailty.

Building on the existing literature [18], future research should focus on designing multi-component interventions, such as combining physical exercise, behavioural modifications, nutritional support, and social engagement, while applying additional frailty indices to further assess their effectiveness to prevent progression of frailty.

In conclusion, this study documents reductions in the level of frailty from baseline to 3 months among older adults with mobility limitations following two 10-week interventions designed to enhance outdoor walking. No significant differences were found between the supervised outdoor walking and telephone-based reminder groups. However, the similar improvement in frailty scores coupled with distinct changes in the underlying frailty indicators highlights the value of employing diverse intervention strategies to address frailty. This study offers two actionable approaches for consideration and underscores the need for broader, innovative paradigms to reduce frailty in the growing older population.

## Data Availability

Data cannot be made publicly available due to ethical and legal restrictions. Specifically, permission for public data sharing was not obtained from the Research Ethics Boards at the University of Alberta, University of Manitoba, University of Toronto, or McGill University, nor was consent for public access obtained from participants. However, data are available upon reasonable request and subject to review by the appropriate Research Ethics Board. Requests for access to the data can be directed to the corresponding author or the Health Sciences Research Ethics Board at the University of Toronto.

## Acknowledgements

We thank all participants for generously contributing their time and engagement to this study. We gratefully acknowledge the community-based organizations and parks in Winnipeg (Assiniboine Park, Kildonan Park, St. Vital Park), Edmonton (Emily Murphy Park, William Hawrelak Park), Toronto (Better Living Centre, Edwards Gardens), and Montreal (Centennial Hall/Memorial Park), where workshops and outdoor walk group sessions were conducted. We thank the study personnel and students who supported the study.

## Supporting information

**S1 Fig. Frailty sum score within and after pooling intervention groups at each evaluation time point**

**S2 Fig. Change in individual frailty phenotype across study time points in the pooled sample of GO-OUT participants**

**S1 Table. Summary of missing frailty scores at each study time point**

**S2 Table. Baseline characteristics of participants who remained in the GO-OUT study and those who withdrew the subsequent evaluation time points (3 or 5.5 months)**

**S3 Table. Presence of the five frailty indicators by intervention group and time point**

**S4 Table. Results of mixed-effects logistic regression models of change in frailty over time in the pooled sample of GO-OUT participants (sensitivity analysis)**

## Notes

### Competing Interest Statement

The authors have declared no competing interest.

### Clinical Trial

NCT03292510

### Clinical Protocols

https://pubmed.ncbi.nlm.nih.gov/31005945/

### Author Declarations

The Research Ethics Board at the University of Toronto approved the trial protocol and this secondary data analysis (Protocol # 00035251).

